# Diagnostic accuracy and utility of SARS-CoV-2 antigen lateral flow assays in medical admissions with possible COVID-19

**DOI:** 10.1101/2021.01.27.21250428

**Authors:** Hamish Houston, Ankur Gupta-Wright, Edward Toke-Bjolgerud, James Biggin-Lamming, Laurence John

## Abstract

We evaluated diagnostic accuracy of the Innova SARS-CoV-2 Antigen Rapid Qualitative Test compared to SARS-CoV-2 RT-PCR from nasopharyngeal swabs in adult admissions who met the COVID-19 case definition at a busy acute hospital in the UK. We found the Innova SARS-CoV-2 Antigen Rapid Qualitative Test had a good specificity in patients with symptoms of COVID-19 presenting to hospital. The Innova LFA can be used to rapidly identify COVID-19 cases amongst hospital admissions meeting the COVID-19 case definition, allowing patients to be allocated to COVID-19 cohort areas.

## Text

The scale-up of SARS-CoV-2 antigen lateral flow assays (LFAs) has caused much controversy, with concerns about lower sensitivity in asymptomatic individuals and when assays are performed by operators without healthcare training.^1,2^ The proposed benefits of SARS-CoV-2 antigen LFAs are high specificity, fast turnaround times for results (under 30 minutes) and ease of scalability.^3^ These assays are of potential utility for rapidly identifying SARS-CoV-2 in patients who fit the COVID-19 case definition and require hospital admission as prompt isolation prevents nosocomial transmission. Isolation rooms are often limited and capacity easily overwhelmed, necessitating the cohorting of patients with proven COVID-19. Even using rapid platforms, SARS-CoV-2 RT-PCR turnaround times are often too slow to inform patient placement from emergency departments (EDs).^4^ SARS-CoV-2 LFAs could help improve flow of patients from the ED into ‘COVID-19 positive’ cohorts and reduce pressure on limited hospital isolation rooms. However, little data exists on their diagnostic accuracy in this setting.

We therefore evaluated diagnostic accuracy of the Innova SARS-CoV-2 Antigen Rapid Qualitative Test (Lotus Global Company, London, UK) compared to SARS-CoV-2 RT-PCR from nasopharyngeal swabs (NPS) in adult admissions who met the COVID-19 case definition at a busy acute hospital in the UK.^5^ The Innova LFA was performed as per the manufacturer’s instructions by appropriately trained health-care assistants in the ED. A second NPS was simultaneously sent for SARS-CoV-2 RT-PCR. Between the 17^th^ November 2020 and 31^st^ December 2020, 728 patients presenting to the ED met the COVID-19 case definition and had valid Innova LFA and RT-PCR results. Baseline characteristics are shown in Table 1A. 55·1% were male and median age was 67·5 years. 264 patients tested positive by Innova LFA. Those testing positive were younger (median age 65 vs 71, p=0.038), more unwell (NEWS of 5 vs 3, p<0.001) and more often febrile on arrival (Temperature >38°C in 41·9% vs 15·8%, p<0.001) than those with negative LFA results. Overall, admission SARS-CoV-2 RT-PCR was positive in 38·5% (280/728).

**Table 1:** Baseline Characteristics and Diagnostic Performance.

| A – Baseline Characteristics | LFA Negative | LFA Positive | Total | p-value |
| --- | --- | --- | --- | --- |
| N | 464 | 264 | 728 |  |
| Age on arrival (years) median (IQR) | 71 (53.5, 83) (n=464) | 65 (49.5, 80) (n=264) | 67.5 (52, 82) (n=728) | 0.038 |
| Age over 65 years, n (%; 95%CI) | 260 (56.0%, 51.5 - 60.6) | 125 (47.3%, 41.3 - 53.4) | 385 (52.9%, 49.3 - 56.5) | 0.024 |
| Female Sex, n (%) | 211 (45.5%, 40.9 - 50.0) | 116 (43.9%, 38.0 - 49.9) | 327 (44.9%, 41.3 - 48.5) | 0.69 |
| Male Sex, n (%) | 253 (54.5%, 50.0 - 59.1) | 148 (56.1%, 50.1 - 62.0) | 401 (55.1%, 51.5 - 59.7) |  |
| NEWS, median (IQR) | 3 (1, 6) (n=422) | 5 (3, 7) (n=230) | 4 (2, 6) (n=652) | <0.001 |
| Pulse, median (IQR) | 94 (82, 111) (n=426) | 96 (84, 108) (n=229) | 95 (82, 110) (n=655) | 0.66 |
| Systolic BP, median (IQR) | 136 (120, 151) (n=421) | 135.5 (122.5, 149) (n=228) | 136 (121, 151) (n=649) | 0.93 |
| Diastolic BP, median (IQR) | 78 (68, 88) (n=421) | 80 (71, 89) (n=228) | 79 (70, 88) (n=649) | 0.10 |
| Respiratory rate, median (IQR) | 20 (18, 27) (n=425) | 24 (20, 32) (n=228) | 22 (18, 28) (n=653) | <0.001 |
| SpO2 <94%, n (%; 95%CI) | 55 (12.9%, 9.8 - 16.1) | 68 (29.7%, 23.8 - 35.6) | 123 (18.8%, 15.8 - 21.8) | <0.001 |
| Required Supplemental Oxygen, n (%; 95%CI) | 72 (16.9%, 13.3 - 20.4) | 69 (29.9%, 24.0 - 35.8) | 141 (21.4%, 18.3 - 24.6) | <0.001 |
| Temperature >38°C, n (%; 95%CI) | 67 (15.8%, 12.3 - 19.2) | 96 (41.9%, 35.5 - 48.3) | 163 (24.9%, 21.6 - 28.2) | <0.001 |
| B – Diagnostic Performance | LFA Negative | LFA Positive | Total |  |
| N | 464 | 264 | 728 |  |
| SARS-CoV2 RNA Detectable on RT-PCR, n (%) | 38 | 242 | 280 | Sensitivity = 86.4% (95%CI 81.9-90.0) |
| SARS-CoV2 RNA Undetectable on RT-PCR, n (%) | 426 | 22 | 448 | Specificity = 95.1% (95%CI 92.6-96.7) |
|  | NPV = 91.8% (95%CI 87.7-94.5) | PPV = 91.8% (95%CI 87.7-94.5) |  |  |

**A - Baseline characteristics and SARS-CoV-2 RT-PCR results for patients testing positive and negative by the Innova Lateral Flow Antigen (LFA) Assay.** For observations on arrival, 9.6 to 10.9% of data were missing. Pair-wise comparisons were performed using chi-squared tests for proportions, t-tests for means and Wilcoxon rank sum for median. \*P-values are shown for the comparison between the LFA positive and LFA negative groups. IQR=Inter-quartile range, CI=Confidence Interval, NEWS=National Early Warning Score, SpO2=Oxygen Saturations.
**B - Diagnostic performance of the Innova Lateral Flow Antigen (LFA) Assay compared to a single SARS-CoV-2 RT-PCR from nasopharyngeal swab on admission.** PPV = Positive Predictive Value, NPV = Negative Predictive Value, CI = confidence interval.

Compared to SARS-CoV-2 RT-PCR as the reference standard, the Innova LFA had sensitivity of 86·4% (242/280, 95% Confidence Interval [CI] 81·9-90·0) and specificity of 95·1% (426/448, 95%CI 92·6-96·7) (Table 1B). 22/448 (4·9%) patients with a negative SARS-CoV-2 RT-PCR on admission had a positive LFA. 8 of these 22 patients reported a positive COVID-19 test result up to 14 days prior to admission and 5/22 subsequently had a positive PCR result within 5 days of admission. 13/22 had chest radiograph features consistent with ‘classic/probable COVID-19’ as reported by a radiologist. Only 5/22 patients had no PCR or radiological evidence of COVID-19. 1/5 patients reported a confirmed household contact and only 2/5 left hospital with a diagnosis other than COVID-19. This suggests the lower than expected specificity of Innova LFA is likely to be a result of an imperfect reference standard, and specificity would be higher if using a clinical and RT-PCR based composite reference standard.^6^

38 patients had negative Innova LFAs but positive PCR results. 20/38 had cycle threshold (Ct) values available, with median Ct values of 29 (IQR 27-35). Innova LFA results were available a median 3·2 hours after arrival (IQR 2·0-4·3, n=681) compared to 13·8 hours (IQR 9·9-18·2, n=679) for RT-PCR. 57·1% (n=35) had chest radiographs which were reported as typical for COVID-19. Of those with symptom duration recorded, 77.3% (17/22) were symptomatic for at least 7 days prior to attending the ED.

Accounting for the inadequacy of a single SARS-CoV-2 RT-PCR as a reference standard, we found the Innova SARS-CoV-2 Antigen Rapid Qualitative Test had good specificity in patients with symptoms of COVID-19 presenting to hospital. Sensitivity in this setting was high (86.4%) when compared to pre-clinical evaluation studies.^1^ Furthermore, results were mostly available within a few hours of presentation, allowing transfer of patients to COVID-19 cohort areas and reducing demand for isolation rooms whilst awaiting PCR results. Placing patients in the ‘right bed’ first-time is also likely to reduce delays in care and increase efficiency, and allows isolation rooms to be prioritised for individuals requiring admission with suspected COVID-19 but negative LFA results. Of the 38 patients with COVID-19 (based on SARS-CoV-2 RT-PCR) who were ‘missed’ by the Innova LFA, median Ct values were reasonably high, and correspond to viral loads associated with lower sensitivity in previous studies.^1^ However, sensitivity of the Innova LFA appears lower than some other SARS-CoV-2 viral antigen LFAs.^7^ Importantly, individuals requiring admission with suspected COVID-19 should not be moved out of isolation on the basis of a negative SARS-CoV-2 viral antigen LFA results.

In summary, the Innova LFA can be used to rapidly identify COVID-19 cases amongst hospital admissions meeting the COVID-19 case definition with good diagnostic accuracy, and rapidly identify patients that can be allocated to COVID-19 cohort areas. Based on these data, this application of COVID-19 LFAs has now been recommended by NHS England.^8^

## Data Availability

Data and statistical code available upon reasonable request.

## Declarations

### Approval

The study was approved by the London North West University Hospitals Trust Research and Development Committee, and given the SARS-CoV-2 antigen lateral flow assay was implemented as part of routine clinical care and this was a retrospective review using routinely collected clinical data, they deemed formal ethical approval was not required.

## Acknowledgements

We would like to acknowledge to all the clinical staff at Northwick Park Hospital who cared for the patients involved in this study, particularly the point-of-care team within the emergency department.

